# Modeling and Predicting Antibody Durability for mRNA-1273 Vaccine for SARS-CoV-2 Variants

**DOI:** 10.1101/2021.05.04.21256537

**Authors:** George Luo, Zhongbo Hu, John J. Letterio

## Abstract

Recently, the antibody titer levels have been followed in 33 adults who received the mRNA-1273 vaccine for 6 months. With single dose estimated effectiveness of 92.1%, we combine this knowledge with corresponding antibody levels to model and estimate the long-term durability of mRNA-1273 vaccine. Additionally, we integrate studies about differences in antibody neutralization to SARS-CoV-2 variants to understand how variants can affect the durability of the vaccine.

The estimated days after first injection for binding antibodies to fall below levels of those from day 15 is 411 days. The estimated days after first injection to fall below the lower limit of detection of 20 GMTs is 327 days for pseudovirus neutralization and 461 days for live virus neutralization. Our model has pseudovirus neutralization against variant B.1.351 falling below 20 GMT on day 100; variant P.1 on day 202, variant B.1.429 on day 258; and variant B.1.1.7 on day 309.

The data used contains many limitations including the small sample size with older age bias, sensitivity of the neutralization assays, and limited data on variants. Still, we believe mRNA-1273 two dose vaccine can provide over a year of protection against COVID-19 from the initial D614G variant. It is likely by the second year, protection against COVID-19 will fall below single dose efficacy. Therefore, there should be consideration for a booster shot a year after the first set of vaccines. If there is an observed increase in variants with higher resistance such as B.1.351 and P.1, a booster vaccine against the newer variants should be considered to increase protection against resistant variants.

## Background

The severe acute respiratory syndrome coronavirus 2 (SARS-CoV-2) emerged in 2019, and its spread throughout the world has caused the coronavirus disease 2019 (COVID-19) pandemic^1^. COVID-19 mRNA-1273 vaccine can prevent symptomatic COVID-19 illness by about 94%^2^ but its durability of protection to COVID-19 is unknown. Recently, the antibody titer levels have been followed in 33 adults who received the mRNA-1273 vaccine for 6 months.^3^ With single dose vaccine estimated efficacy of 92.1%,^4^ we combine this knowledge with antibody titer and neutralization levels to estimate the long-term durability of mRNA-1273 vaccine. Additionally, we integrate studies about differences in antibody neutralization to SARS-CoV-2 variants to understand how variants can affect the durability of the vaccine.^5,6^

## Results

We modeled data using a power-law regression for binding antibodies, measured by an enzyme-linked immunosorbent assay against SARS-CoV-2 spike receptor–binding domain, and exponential decay for psuedovirus and live virus neutralization assays (Figure 1A-C)^3^. We utilized data from day 43 after first injection and beyond to create regression models for the binding antibodies, pseudovirus neutralization, and live virus neutralization (R^2^ values of 0.9917, 0.9972, and 0.9872 respectively, see Supplementary Appendix).^3^ The estimated days after first injection for binding antibodies to fall below levels of those from day 15 is 411 days (95% CI, 304 to 519). Both the pseudovirus and live virus neutralization assays have a lower limit of detection of 20 for the 50% inhibitory dilutions with the estimated day 15 and 29 geometric mean end-point titers (GMTs) in the vicinity of this threshold. Therefore, it is possible that these assays were not sensitive enough to detect lower levels of antibodies that still provide protection to COVID-19 infections. The estimated days after first injection to fall below the lower limit of detection of 20 GMTs is 327 days (95% CI, 307 to 372) for pseudovirus neutralization and 461 days (95% CI, 416 to 512) for live virus neutralization. There appears to be a trend of longer durability with lower age but because of small number of samples, this was inconclusive.

**Figure 1.**
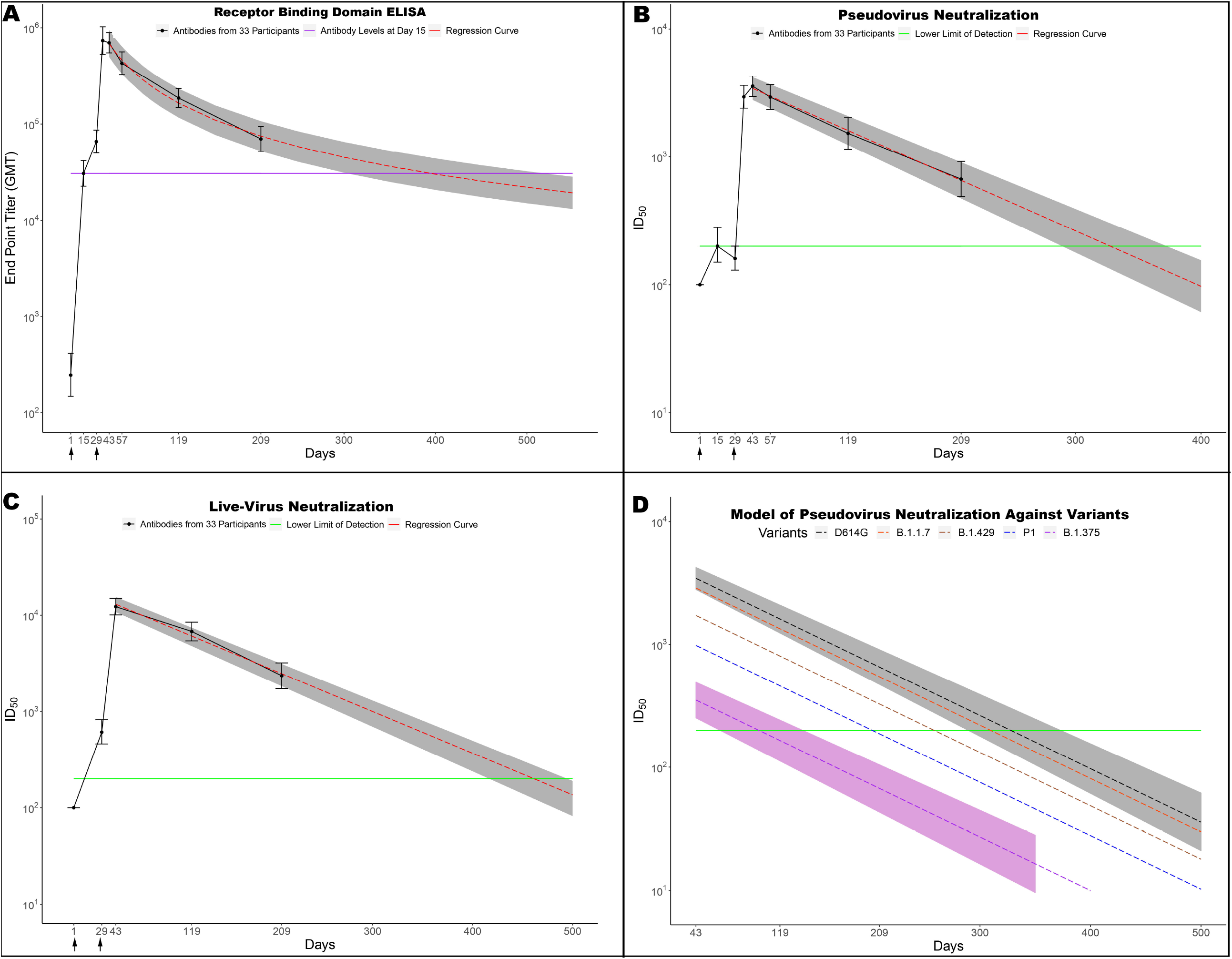
Prediction of Antibody Binding and Neutralization to SARS-CoV2 and Variants over Time. 33 participants were injected with 100 ug of mRNA-1273 on day 1 and 29. Serum samples were collected up to day 209 and tested in assays. The titer for binding to receptor binding domain was assessed by enzyme-linked immunosorbent assay (ELISA) and modeled to best fit the data for day 43 and beyond (Panel A). The solid purple line represents the average antibodies levels on day 15 while the red line is regression curve. The models for 50% inhibitory dilution (ID_50_) titer by pseudovirus neutralization (Panel B) and by live virus neutralization (Panel C) are shown. Pseudovirus neutralization is modeled for variants of SARS-CoV2 (panel D) with prototypical D614G (black), B1.1.7 (orange), B.1.429 (brown), P.1 (blue), and B.1.351 (violet). The solid green line represents the limit of detection for the neutralization assays. I bars are 95% confidence intervals. The shaded gray area represents the predicted 95% confidence interval for regression curves for the 33 participants while the shaded violet area represents the predicted 95% confidence interval for regression curves of B.1.351 for the 33 participants.

We estimated differences in virus neutralization of the prototypical D614G variant to variants B.1.351 and B.1.429. with data from X. Shen et. al.^5^ and neutralization of D614G variant to variants B.1.1.7 and P.1 with data from K. Wu. Et. al.^6^ Our model has pseudovirus neutralization against variant B.1.351 falling below 20 GMT on day 100 (95% CI, 65-139); variant P.1 on day 202, variant B.1.429 on day 258 (95% CI, 213 – 306); and variant B.1.1.7 on day 309 (Figure 1D). There was insufficient data on variants P.1 and B.1.1.7 to calculate the variability and a confidence interval. Our model suggests that the two-dose vaccine will provide some protection for all above variants but considerably lower durability and higher variability for variants B.1.351 and probably for P.1. Similar neutralization results have been found in other studies that single dose of mRNA vaccine was largely ineffective to induce neutralization against B.1.351, but that two doses could achieve neutralization^7^.

## Conclusion and Discussion

The data contains many limitations including small sample size with a bias towards older age, low sensitivity of the neutralization assays, and limited data on variants differences. Still, we believe mRNA-1273 two dose vaccine can provide over a year of protection against COVID-19 from the initial D614G variant. It is likely that by the second year, antibody protection against COVID-19 will fall below single dose efficacy although memory B cells may still protect individuals^7^. Therefore, there should be consideration for a booster shot a year after the first set of vaccines. If there is an observed increase in variants with higher resistance such as B.1.351 and P.1 in the population of vaccination, a booster or vaccine against the newer variants should be considered to increase protection against resistant variants.

## Data Availability

Data is available through supplemental document.

