## Supplemental Appendix for "Modeling and Predicting Antibody Durability for mRNA-1273 Vaccine for SARS-CoV-2 Variants"

**Supplemental Methods**

**Antibody Data**. Data was used from supplemental tables in N. Doria-Rose et. al. 2020 for the 33 participants who received the mRNA-1273 vaccine[1]. Data from X. shen et al. and K Wu. et. al. was used to estimate the variant ID50 compared to the prototypical D614G[2,3].

**Statistic Methods**. To model the antibody binding and neutralization, power law and exponential decay regression was used, respectively. Aggregate data for all regressions was used from study day 43 and later after injection. To calculate a regression curve for antibody binding, $\log_{10}$ transformation was applied to both titer and study day:

$$log_{10}(Titer) = \alpha + \beta * log_{10}\,(\text{study day}_i) + e_i$$

where $\alpha$ is intercept and $\beta$ is decay rate. The study day is days after first injection with the day of injection being day 1. However, the data used for the model starts on day 43 so days before study day 43 is not part of our model and estimates. $e_I$ is the error of each point relative to the regression. Using this equation, the coefficient of determination ($R^2$) and confidence interval were calculated with linear regression in R. If the calculated confidence intervals at the study date (43,57,119,209) were less than the confidence intervals of the original data or did not fit the regression curve ($p > 0.05$), an alternative confidence interval was calculated using the following equation:

$$log_{10}(Titer_{CI2}) = \alpha_{CI2} + \beta_{CI2} * log_{10}\,(\text{study day}_i) + e_{CI2}$$

where $a_{ci2}$ is intercept and $b_{ci2}$ is decay rate. $Titer_{CI2}$ is the titer of the lower and upper bound of the confidence interval of the original data and $e_{CI}$ is the error of each point relative to the regression.

For neutralization assays, an exponential decay regression was used with $\log_{10}$ transformation was applied to the $ID_{50}$ titer levels resulting in the following equation:

$$log_{10}(Titer) = \alpha + \beta * \text{study day}_i + e_i$$

where $\alpha$ is intercept and $\beta$ is decay rate. $e_i$ is the error of each point relative to the regression. Using this equation, the coefficient of determination ($R^2$) and confidence interval were calculated. If the calculated confidence intervals at the study date (43,57,119,209) were less than the confidence intervals of the original data or did not fit the regression curve ($p > 0.05$), an alternative confidence interval was calculated using the following equation:

$$log_{10}(Titer_{CI2}) = \alpha_{CI2} + \beta_{CI2} * \text{study day}_i + e_{CI2}$$

where $a_{ci2}$ is intercept and $b_{ci2}$ is decay rate. $Titer_{CI2}$ is the titer of the lower and upper bound of the confidence interval of the original data and $e_{CI}$ is the error of each point relative to the regression.

To calculate variant $ID_{50}$, the correlation between the neutralization $ID_{50}$ of D614G and the ratio of $ID_{50}$ of D614G/$ID_{50}$ of variant was first investigated[2]. For variants B.1.351 and B.1.429, the $R^2$ of 0.0039 and 0.0042 were observed respectively ($p > 0.32$ and $p > 0.31$) so we assumed that neutralization levels of D614G and the neutralization ratio of variant/D614G were independent. Data from X. Shen. et. al. was used to estimate the $ID_{50}$ of 614G / $ID_{50}$ of B.1.351 and B.1.429[2]. Data from K. Wu. et. al. was used to estimate the $ID_{50}$ of 614G / $ID_{50}$ of B.1.1.7 and P.1[3]. To calculate the mean $ID_{50}$ of a variant at study $day_i$, the following equation was used:

$$log_{10}(Titer_V) * m_v = \alpha + \beta * (\text{study day}_i) + e_i$$

where $\alpha$, $\beta$, and $e_i$ are the same as previous equations and $m_v$ is the ratio of the $ID_{50}$ of D614G/$ID_{50}$ of variant. This allowed calculation of the estimated geometric mean titer of the $ID_{50}$ for any of the variant on study day *i*. This equation was used to plot the variant regression curves for figure 1D.

The $log_{10}$ transformed standard deviation of the $ID_{50}$ of the original 33 participants and the standard deviation of the $ID_{50}$ of D614G/$ID_{50}$ of B.1.351. were used to generated a combined standard deviation through the following equation:

$$SD_C = \sqrt{(SD_P)^2 + (SD_V)^2}$$

where $SD_c$ is the $log_{10}$ of the combined standard deviation. $SD_P$ is the $log_{10}$ of the standard deviation of the pseudovirus neutralization $ID_{50}$ of the 33 participants while the $SD_v$ is the $log_{10}$ of the standard deviation of the $ID_{50}$ of D614G / $ID_{50}$ of variant B.1.351. Using the $SD_c$, a confidence interval for the $ID_{50}$ of B.1.351 and B.1.429 of the 33 participants was estimated for days 43,57,109, and 209.

**Supplemental Table 1. Antibody Binding and Viral Neutralization Regression Calculations**

| | RBD ELISA | | Pseudovirus Neutralization | | Live-Virus Neutralization | |
|---|---|---|---|---|---|---|
| Variables | Value | p-value | Value | p-value | Value | p-value |
| $\alpha$ (intercept) | 8.130 | 0.0005 | 2.723 | <0.0001 | 3.300 | 0.01 |
| $\beta$ (decay rate) | -1.404 | 0.004 | -0.004339 | 0.001 | -0.004335 | 0.07 |
| $R^2$ | 0.9917 | 0.004 | 0.9971 | 0.001 | 0.9872 | 0.07 |
| $\alpha_{CI}$ | (8.033 to 8.228) | 0.0002 | (2.672 to 2.773) | < 0.0001 | (2.7382 to 3.8628) | 0.055 |
| $\beta_{CI}$ | (-1.429 to 1.378) | 0.0002 | (-0.004471 to - 0.004207) | 0.0008 | (-0.00455 to - 0.00412) | 0.25 |
| $R^2{}_{CI}$ | 0.9958 | 0.002 | 0.9983 | 0.0008 | 0.8548 | 0.25 |
| $\alpha_{CI2}$ | not calculated | n/a | (2.646 to 2.797) | 0.0001 | 3.2324 to 3.3689 | 0.01 |
| $\beta_{CI2}$ | not calculated | n/a | (-0.004648 to - 0.004015) | 0.003 | (-0.004639 to - 0.004038) | 0.07 |
| $R^2{}_{CI2}$ | not calculated | n/a | 0.994 | 0.003 | 0.985 | 0.07 |

RBD ELISA utilized confidence interval calculation from regression modeling so the alternative confidence interval was not calculated. Pseudovirus neutralization and Live-virus neutralization used alternative confidence interval for modeling.

**Table 2. Variant Pseudovirus Neutralization Modeling Calculations**

| | B.1.351 | B.1.429 | B.1.1.7 | P.1 |
|---|---|---|---|---|
| $m_v$ (ID50 D614G / ID50 Variant) | 9.75 | 2.04 | 1.2 | 3.5 |
| $SD_V$ | 0.246 | 0.224 | NE | NE |
| $SD_P$ on Day 43 | 0.227 | 0.227 | 0.227 | 0.227 |
| $SD_P$ on Day 57 | 0.275 | 0.275 | 0.275 | 0.275 |
| $SD_P$ on Day 109 | 0.348 | 0.348 | 0.348 | 0.348 |
| $SD_P$ on Day 209 | 0.386 | 0.386 | 0.386 | 0.386 |
| $SD_C$ on Day 43 | 0.335 | 0.318 | NE | NE |
| $SD_C$ on Day 57 | 0.369 | 0.354 | NE | NE |
| $SD_C$ on Day 109 | 0.426 | 0.414 | NE | NE |
| $SD_C$ on Day 209 | 0.458 | 0.446 | NE | NE |
| 95% CI on Day 43 | 26.8 to 50.3 | 129.7 to 236.4 | NE | NE |
| 95% CI on Day 57 | 21.3 to 42.7 | 103.0 to 200.8 | NE | NE |
| 95% CI on Day 109 | 10.5 to 23.5 | 50.7 to 110.5 | NE | NE |
| 95% CI on Day 209 | 4.5 to 10.6 | 21.5 to 49.9 | NE | NE |

NE= not estimated due to lack of data
